# The MINIFLOW Study: Evaluating Non-Invasive Imaging Modalities for Follow-Up of Flow Diverter Stent Treated Cerebral Aneurysms

**DOI:** 10.1101/2025.09.26.25336776

**Authors:** Viktoria E. Shimanskaya, Maud van Elderen, Sjoert A.H. Pegge, Bram A.C.M. Fasen, Elle Vermeulen, Bart A.J.M. Wagemans, Marlien W. Aalbers, Joost de Vries, Frederick J.A. Meijer, Hieronymus D. Boogaarts

**Affiliations:** Department of Neurosurgery, Radboud University Medical Center, Nijmegen, The Netherlands; Department of Medical Imaging, Radboud University Medical Center, Nijmegen, The Netherlands; Department of Neurosurgery, University Hospital Brussel, Brussels, Belgium; Department of Radiology, Maastricht University Medical Center, Maastricht, The Netherlands; Department of Ophthalmology, University Medical Center Groningen, Groningen, The Netherlands

## Abstract

**BACKGROUND:** This study aimed to evaluate subtraction CT angiography (sCTA) and MR angiography (MRA) as potential non-invasive alternatives for digital subtraction angiography (DSA) for the follow-up of intracranial aneurysms treated with flow diverter stent (FDS).

**METHODS:** Forty patients with intracranial aneurysms treated with FDS were enrolled between August 2019 and November 2024 to evaluate the diagnostic performance of sCTA and MRA in assessment of aneurysm occlusion after FDS treatment. All patients were scheduled to undergo sCTA, MRA and DSA within 24 hours. sCTA was performed on a 160-row detector ultra-high resolution CT scanner. TOF MRA pre- and post-contrast, DWI and T1 sequences were acquired using a 3T MRI scanner. All studies were assessed by two blinded observers. Sensitivity, specificity, negative and positive predictive values were calculated for aneurysm occlusion. Interrater variability was assessed using the Cohen’s kappa.

**RESULTS:** Compared to DSA, specificity for evaluation of complete aneurysm occlusion was high for sCTA (88%), TOF MRA (92%) and post-contrast TOF MRA (83%). Post-contrast TOF MRA had the highest sensitivity (93%) and negative predictive value (95%). Interrater variability was good to excellent for all three modalities. Complications associated with DSA were observed in five patients; in one case, a 24-hour hospital admission was necessary for observation.

**CONCLUSION:** sCTA, pre- and post-contrast TOF MRA seem to be appropriate for evaluating aneurysmal occlusion after FDS treatment, considering the high specificity and negative predictive value and non-invasive nature.

## INTRODUCTION

A flow diverter stent (FDS) is a stent-like device aimed for endoluminal parent vessel reconstruction and flow modification for treatment of cerebral aneurysms, especially for those aneurysms with a configuration unfavorable for coiling.^1–3^ Currently, digital subtraction angiography (DSA) is considered the gold standard for assessment of aneurysm occlusion after FDS treatment. However, DSA is an invasive imaging technique with a risk of vascular and thromboembolic complications.^4–6^ In addition, it does not depict the surroundings of the aneurysm, thrombosed aneurysmal sac or brain parenchyma, which are of particular interest after treatment with FDS.

This has led to a growing interest in alternative non-invasive imaging modalities with similar diagnostic accuracy, such as subtraction CT angiography (sCTA) and MR angiography (MRA). Various publications report the use of sCTA and MRA as a valuable imaging modality for follow up after FDS.^7–9^ However, the quality of these reports is limited by small numbers of included patients, inhomogeneous study populations in terms of aneurysm characteristics and treatment modalities, and even limited availability of DSA as a reference. The aim of this study is to determine whether sCTA or MRA could substitute DSA for assessment of aneurysm occlusion after FDS treatment.

## METHODS

### Study cohort

A prospective cohort study was performed at Radboud University Medical Center, Nijmegen. Approval was granted by the regional medical-ethical committee, and all patients provided written informed consent. This study was registered on a Dutch trial registry https://onderzoekmetmensen.nl/en under identifier NL-OMON47976. Patients treated with FDS for intracranial aneurysms were consecutively enrolled between August 2019 and December 2024. Inclusion was based on identification by their treating physician, either at time of treatment or at follow-up visits. Exclusion criteria were as follows: patients under 18 years of age, pregnancy, known allergies or contraindications to the used contrast agents, renal insufficiency (MDRD < 60 mL/min/1.73 m2) and use of nephrotoxic medications. Additionally, patients who were involved in other studies with outcomes potentially influenced by the additional imaging performed in this study were excluded. All participants were scheduled to undergo sCTA, DSA, and MRI, with all scans conducted within 24 hours to minimize variability in outcome. The order of the investigations was random depending on the available timeslot with MRI performed as the final imaging modality. Patients with contra-indications for MRI underwent sCTA and DSA only.

The aim of this study was to show non-inferiority of sCTA and TOF MRA compared to DSA in assessing complete aneurysm occlusion. The estimated chance for a false negative result in assessment of untreated intracranial aneurysms is 1.0-1.75% based on the available data on sCTA.^10^ Taking into account a type I error of 0.05 for a one-sided test, a power of 80%, a chance of 1.0% and a margin of 12% for a false negative result, this study required at least 38 patients with at least 24 completely occluded aneurysms, allowing a maximum of one false negative result. To ensure that enough patients could be included in the final analysis, 40 patients were recruited.

This manuscript was prepared in accordance with the STARD reporting guidelines for diagnostic accuracy studies.^11^

### Imaging acquisition and analysis

#### DSA

DSA was performed by an experienced neurosurgeon using a flat-panel C-arm angiography system (Allura Xper FD20; Philips healthcare, Best, the Netherlands). Catheterization was performed via either the femoral or radial artery, with ultrasound guidance. A 6 French sheath was used for vascular access. Catheters included Sim vertebral catheters (Terumo; 2.0–4.0 French) and the Vitek catheter (Terumo; 5 French). During 3D rotational angiographic acquisition using a cerebral soft-tissue protocol (XperCT; Philips Healthcare), a selective contrast bolus was delivered via power injector (Angiomat Illumena; Covidien/Mallinckrodt, Lake Forest, California). Images were analyzed on a dedicated workstation (Xtra Vision; Philips Healthcare) to determine optimal 2D projections for stent evaluation. The arterial puncture site closure was achieved using an Angio-Seal, TR band, or manual compression. The average contrast volume used was 82 mL (iomeprol 300 mg iodine/mL; Iomeron, Bracco, Milan, Italy). The estimated radiation dose for the complete DSA protocol was approximately 3.0 mSv.

#### sCTA

sCTA scans were performed using a 160-row detector ultra-high resolution CT scanner (Aquilion Precision, Canon Medical Systems Corporation, Otawara, Japan), as described in a previous study.^12^ The protocol included a non-enhanced acquisition at 120 kV and 75 mAs, which was followed by a contrast-enhanced acquisition at 120 kV and 150 mAs, for which 60 mL of Iomeron 300 contrast was administered intravenously. Using bolus-tracking, scanning time after contrast injection was determined. Images were acquired with a slice thickness of 0.25mm, a matrix of 1024×1024, and a pitch factor of 0.569. The estimated radiation dose was 2.45 mSv. Single-energy metal artifact reduction (SEMAR) and subtraction algorithms were applied.

#### TOF MRA

MRA scans were acquired using a 3T scanner (Magnetom Skyra scanner, Siemens Healthineers, Erlangen, Germany). A time-of-flight MRA sequence (TR 24 ms; TE 3.93 ms; slice thickness 1 mm; flip angle 8°; and matrix of 224×224) was acquired before and after intravenous injection of 7.5 mmol Dotarem. Additionally, diffusion-weighted imaging (DWI) (TR 5660 ms; TE 64 ms; slice thickness 5 mm; flip angle 180°; and matrix of 160×160) and a 3D T1 sequences were obtained (TR 2200 ms; TE 2.43 ms; slice thickness 1 mm; flip angle 8°; and matrix 224×224).

### Image analysis

All imaging studies were independently reviewed by two blinded observers. Both the sCTA and MRA scans were analyzed by two neuroradiologists (SP and BF), with a two-week interval and in random order to prevent recognition bias. DSA images were assessed by two neurointerventionalists (EV and BW). In case of discrepancies between observers a final decision was rendered by consensus. None of the readers were involved in the clinical treatment of the patients. The readers were provided with limited clinical information and included the number, location and shape of the aneurysm(s); aneurysm size; the type of FDS used; details of any additional treatments, such as coiling or clipping.

All imaging modalities were analyzed on a dedicated clinical workstation (Impax, version 8.2.0, Agfa Healthcare). Image quality and the presence of artifacts were systematically assessed for all scans. Scores for image quality ranged from 1 (non-diagnostic) to 5 (excellent quality) on Likert scale while the presence of artifacts was scored on a scale from 1 (severe artefacts) to 5 (no artefacts).^13^ The primary outcome, aneurysm occlusion, was measured using the Raymond-Roy (RR) classification.^14^ Grade 1 was defined as complete occlusion, Grade 2 as a neck residual, and Grade 3 as aneurysm remnant. A subgroup analysis of the primary outcome measure was conducted in patients without coils in close proximity to the FDS. Secondary outcomes included wall apposition, neck coverage, side branches patency and procedure-related complications.

### Statistical analysis

Statistical analyses were conducted using SPSS (version 29.0; IBM, Armonk, New York). Sensitivity, specificity, positive predictive value (PPV), and negative predictive value (NPV) were calculated for assessing aneurysm occlusion, with DSA serving as the reference modality. One-sided 95% confidence intervals (lower bounds) were calculated using the Clopper–Pearson exact method, based on a type I error of 0.05. Interrater variability was assessed using Cohen’s method to categorize the κ-values as follows: κ < 0.4 indicating positive but poor agreement, κ 0.41-0.75 indicating good agreement, and κ > 0.75 indicating excellent agreement.^15^ Descriptive statistics were applied to summarize patient and aneurysm characteristics.

## RESULTS

Between August 2019 and December 2024, 67 patients were screened for eligibility and 40 patients with 43 aneurysms treated with a FDS were eventually enrolled in this study. Reasons for exclusion were lack of informed consent (n=18), lost to follow-up (n=6), renal insufficiency (n=2), allergy to contrast agent (n=1). The median follow-up time was 9.5 months (range 1-149 months). See table 1 for demographic and clinical characteristics of the included patients.

**Table 1.** patient characteristics.

|  |  |
| --- | --- |
| **Number of patients** | 40 |
| **Gender (n (%))**<br>Female | 31 (77.5%) |
| **Age in years at follow-up (mean (range))** | 55 (26 – 78) |
| **Follow-up time after FDS placement in months (median (range))** | 9.5 (1 – 149) |
| **Total number of aneurysms** | 43 |
| **Number of aneurysms per patient (n (%))**<br>1 aneurysm<br>2 aneurysms<br>3 aneurysms | 38 (95%)<br>1 (2.5%)<br>1 (2.5%) |
| **Aneurysm location (n (%))**<br>Internal carotid artery<br>Posterior communicating artery<br>Middle cerebral artery<br>Superior cerebellar artery<br>Anterior communicating artery<br>Anterior cerebral artery<br>Basilar artery<br>Vertebral artery<br>Posterior inferior cerebellar artery | 22 (51.2%)<br>8 (18.6%)<br>4 (9.3%)<br>3 (7.0%)<br>2 (4.7%)<br>1 (2.3%)<br>1 (2.3%)<br>1 (2.3%)<br>1 (2.3%) |
| **Aneurysm size (n (%))**<br>< 5 mm<br>5 – 10 mm<br>> 10 mm | 11 (25.6%)<br>18 (41.9%)<br>14 (32.6%) |
| **Shape of FDS treated aneurysm (n (%))**<br>Saccular<br>Fusiform<br>Blister<br>Dissecting<br>Other | 38 (88.4%)<br>1 (2.3%)<br>1 (2.3%)<br>1 (2.3%)<br>2 (4.7%) |
| **Types of FDS used**<br>Surpass Evolve<br>Silk vista<br>Derivo<br>Unknown | 35 (87.5%)<br>3 (7.5%)<br>1 (2.5%)<br>1 (2.5%) |
| **Coiling in addition to FDS (n (%))** | 18 (41.9%) |
| **Hypertension (n (%))** | 19 (47.5%) |
| **SAH in medical history (n (%))** | 16 (40.0%) |

FDS = flow diverter stent; SAH = subarachnoid hemorrhage

The mean age of the patients was 55 years (range 26-78), 78% were female. Thirty-seven aneurysms were located in the anterior circulation and six aneurysms in the posterior circulation. Eleven aneurysms were <5 mm, eighteen aneurysms were 5-10 mm and fourteen aneurysms >10 mm. Sixteen patients had subarachnoid hemorrhage in their medical history and eighteen assessed aneurysms had coils in proximity to FDS. In most cases a Surpass Evolve FDS was used (88%), followed by Silk Vista FDS (8%) and Derivo FDS (3%). In one case the patient was treated at another institution and the type of FDS used could not be determined.

Thirty-six patients underwent all three imaging modalities. Four patients did not undergo MRI either due to a contraindication in three cases, or due to a withdrawal after the DSA in one case. In one patient, the pre-contrast TOF MRA was unavailable.

### Image quality

The mean image quality scores, ranging from 1 (non-diagnostic) to 5 (excellent quality) were averaged between the two observers, with highest scores for sCTA (4.1±0.27), followed by DSA (4.0±0.57), MRA pre-contrast (3.9±0.34) and MRA post-contrast (3.9±0.32). Artifacts levels, evaluated on a scale from 1 (severe artefacts) to 5 (no artifacts), varied across modalities. On average, DSA exhibited the least artifacts (4.2±0.75), followed by sCTA (3.3±0.86), MRA pre-contrast (3.0±0.49) and MRA post-contrast (2.9±0.45). Notably, the administration of contrast in MRA did not influence image quality or artifact levels, as mean scores for pre- and post-contrast MRA remained comparable.

### Primary outcome

For the analysis of the aneurysm occlusion the RR classification was dichotomized in complete (grade 1) and incomplete (grade 2 or 3) occlusion. The results of the analysis are shown in table 2.

**Table 2.** diagnostic performance metrics of sCTA, pre-contrast TOF MRA and post-contrast TOF MRA compared to DSA for assessing complete (RR grade 1) and incomplete occlusion (RR grade 2 and 3).

| Modality (number of aneurysms) | Sensitivity (lower bound of 95% one-sided CI) | Specificity (lower bound of 95% one-sided CI) | PPV (lower bound of 95% one-sided CI) | NPV (lower bound of 95% one-sided CI) | Inter-observer agreement |
| --- | --- | --- | --- | --- | --- |
| sCTA (n=42) | 71% (48%) | 88% (69%) | 80% (52%) | 81% (63%) | 0.847 |
| TOF MRA<br>pre-contrast (n = 38) | 71% (44%) | 92%<br>(74%) | 83%<br>(54%) | 85%<br>(66%) | 0.825 |
| TOF MRA<br>post-contrast (n = 39) | 93% (70%) | 83%<br>(64%) | 78%<br>(56%) | 95%<br>(77%) | 0.652 |

sCTA - subtraction CT angiography, TOF MRA - time-of-flight MR angiography, PPV – positive predictive value, NPV – negative predictive value, CI – confidence interval

In our cohort, DSA demonstrated complete occlusion in 26 out of 43 aneurysms (60.5%). One sCTA was excluded from the analysis due to coil-related artefacts that impeded accurate assessment of aneurysm occlusion. In this case, a correct evaluation was possible with TOF MRA. sCTA correctly identified complete occlusion in 22 out of 25 cases, pre-contrast TOF MRA in 22 out of 24 cases and post-contrast TOF-MRA in 20 out of 24 cases. Figure 1 illustrates complete aneurysm occlusion following FD treatment, as demonstrated by all imaging modalities. Incomplete occlusion was accurately determined by sCTA in 12 out of 17 cases. In 5 cases, sCTA overestimated the degree of occlusion (four neck remnants and one residual aneurysm). Pre-contrast TOF MRA correctly assessed incomplete occlusion in 10 out of 14 cases and missed a neck remnant in 4 cases. Post-contrast TOF MRA and DSA demonstrated concordant findings with DSA regarding incomplete occlusion in 14 out of 15 cases. One neck remnant was missed on post-contrast TOF MRA. Figure 2 depicts this missed remnant on both sCTA and post-contrast TOF MRA. Inter-observer agreement for aneurysm occlusion was excellent for sCTA and pre-contrast TOF MRA (0.847 and 0.825, respectively) and good for post-contrast TOF MRA (0.652). All three modalities demonstrated good agreement with DSA in assessing occlusion using the non-dichotomized Raymond-Roy classification, with kappa values of 0.508 for sCTA, 0.539 for pre-contrast TOF MRA, and 0.730 for post-contrast TOF MRA.

**Figure 1.**
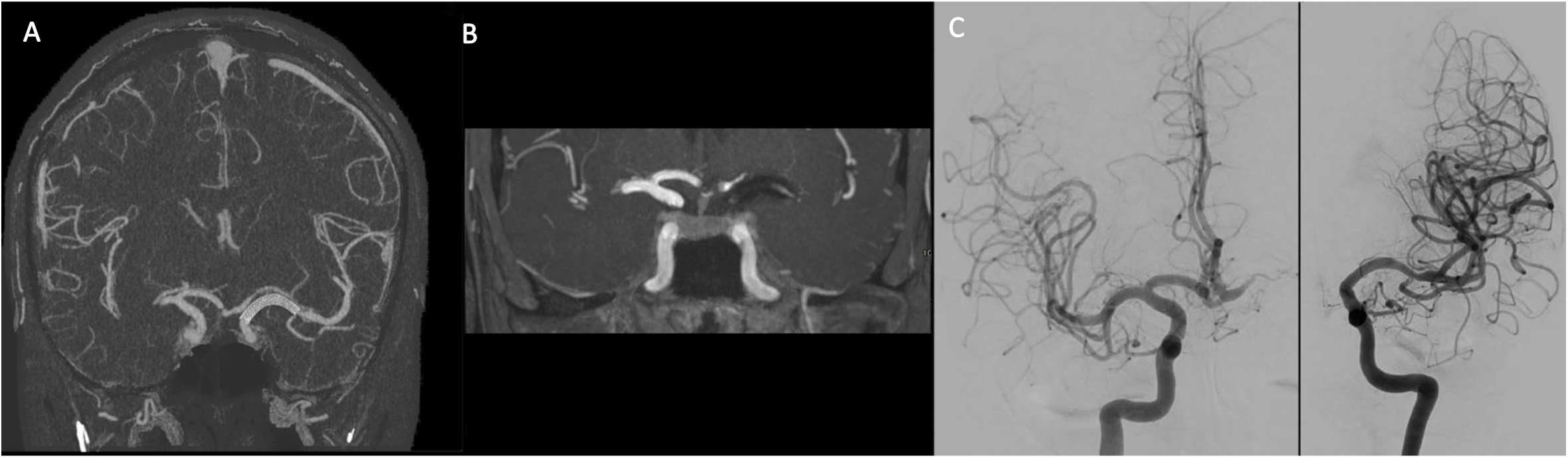
Flow diverter extending from the distal left internal carotid artery to the M1 segment of the left middle cerebral artery (MCA), demonstrating complete occlusion of the left MCA aneurysm on sCTA (A), post-contrast TOF MRA (B), and DSA (C).

**Figure 2.**
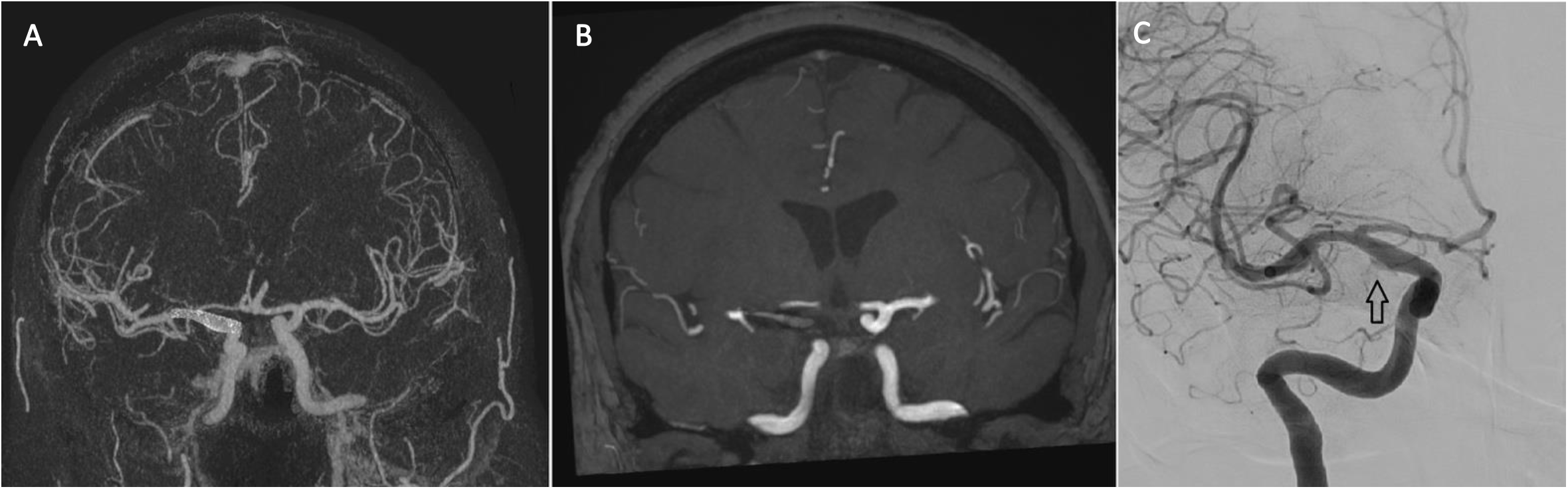
Flow diverter extending from the right internal carotid artery to the M1 segment of the right middle cerebral artery demonstrating complete occlusion on sCTA (A) and post-contrast TOF MRA (B), and a small neck remnant (arrow) on DSA (C).

### Secondary outcome

Wall apposition and neck coverage were assessed across all imaging modalities. In DSA examinations, assessment was feasible in all cases; however, interobserver discrepancies were noted in five cases for wall apposition and in nine cases for neck coverage. For sCTA, both parameters were assessable in all cases, with discrepancies observed in two patients for wall apposition and in one patient for neck coverage. No discrepancies between pre- and post-contrast MRA were found in the assessment of wall apposition or neck coverage. Interobserver discrepancies for pre-and post-contrast MRA were observed in five patients for wall apposition and in four patients for neck coverage. Side branches assessment was feasible in 36 patients on sCTA and in 34 patients on TOF MRA.

### Complications

Complications related to DSA were observed in five cases. Two patients developed a puncture site hematoma, one of whom required hospital admission for 24-hour observation. One patient experienced a transient scotoma without infarction on MRI. Another patient developed transient aphasia during the DSA procedure, suspected to be a contrast-related reaction, as symptoms resolved rapidly post-procedure and follow-up MRI showed no diffusion abnormalities. Additionally, one patient demonstrated a brain area with restricted diffusion on MRI, suggestive of a possible thromboembolic complication related to the DSA. However, the patient remained asymptomatic. No complications were reported following sCTA or MRA.

### Subgroup analyses

A subanalysis was conducted in patients without coils in close proximity to the FDS. The results are summarized in Table 3. Complete aneurysm occlusion on DSA was observed in 15 out of 25 patients. Sensitivity, specificity, PPV and NPV were comparable between sCTA and pre-contrast TOF MRA. Post-contrast TOF MRA demonstrated particularly high specificity, PPV, and NPV. Inter-observer agreement was excellent for sCTA and pre-contrast TOF MRA, with kappa values of 1.0 and 0.814, respectively, and remained good for post-contrast TOF MRA (κ = 0.673).

**Table 3.** diagnostic performance metrics of sCTA, pre-contrast TOF MRA and post-contrast TOF MRA compared to DSA for assessing complete (RR grade 1) and incomplete occlusion (RR grade 2 and 3) for patients without coils.

| Modality (number of aneurysms) | Sensitivity<br>(lower bound of 95% one-sided CI) | Specificity<br>(lower bound of 95% one-sided CI) | PPV<br>(lower bound of 95% one-sided CI) | NPV<br>(lower bound of 95% one-sided CI) | Inter-observer agreement |
| --- | --- | --- | --- | --- | --- |
| sCTA (n=25) | 80%<br>(49%) | 93%<br>(70%) | 89%<br>(56%) | 88%<br>(58%) | 1.0 |
| TOF MRA<br>pre-contrast (n = 24) | 78%<br>(44%) | 93%<br>(70%) | 88%<br>(48%) | 88%<br>(58%) | 0.814 |
| TOF MRA<br>post-contrast (n = 24) | 82%<br>(55%) | 100%<br>(78%) | 100%<br>(69%) | 100%<br>(78%) | 0.673 |

sCTA - subtraction CT angiography, TOF MRA - time-of-flight MR angiography, PPV – positive predictive value, NPV – negative predictive value, CI – confidence interval

## DISCUSSION

This study aimed to evaluate whether sCTA, pre- and post-contrast TOF MRA provide diagnostic accuracy that is non-inferior to DSA in assessing cerebral aneurysm occlusion following FDS treatment, thereby offering a potential non-invasive alternative to DSA. The study findings suggest that these modalities may serve as reliable non-invasive alternatives for follow-up, considering the high specificity and NPV. The analysis specifically aimed to differentiate between complete (RR grade 1) and incomplete aneurysm occlusion (RR grade 2 or 3), given that this distinction holds the greatest clinical significance. Furthermore, the absence of clearly defined cut-off criteria for differentiating between neck residual and aneurysm remnant resulted in considerable variability in the assessment of RR grade 2 and 3 occlusions for sCTA and MRA, and in lesser extent for DSA. Nonetheless, all imaging modalities demonstrated good agreement with DSA in assessing occlusion using the non-dichotomized Raymond-Roy classification.

sCTA demonstrated high specificity (88%) and a relatively high negative predictive value (81%) for neck rest and aneurysm rest, indicating that negative findings—i.e., complete aneurysm occlusion—are generally reliable and may eliminate the need for additional DSA. The positive predictive value for the detection of neck or aneurysm remnant was also high (80%). However, the sensitivity was lower (71%), suggesting that some cases of incomplete occlusion may be missed. Notably, sCTA showed the highest interobserver agreement among the modalities, underscoring its reliability and consistency across different assessors.

A previous pilot study investigated the diagnostic accuracy of sCTA for assessing aneurysm occlusion, reporting both 100% sensitivity and specificity, as well as perfect interobserver agreement.^9^ While our specificity findings are comparable, the sensitivity observed in our study is notably lower. This discrepancy could be explained by differences in study design and patient cohorts: the pilot study included patients with known aneurysm remnants and had a shorter follow-up period (1 month), compared to a median follow-up of 9.5 months in our cohort.

Pre-contrast TOF MRA yielded results comparable to those of sCTA, demonstrating high specificity (92%), relatively high NPV (85%) and comparable sensitivity and PPV (71% and 83%, respectively) suggesting that incomplete occlusions could still be missed, and positive findings might require confirmation with DSA.

In contrast, post-contrast TOF MRA demonstrated high sensitivity (93%) and a good PPV (83%), making it effective in detecting incomplete occlusions. Although specificity was slightly lower (83%), it remained acceptable, and the NPV was high (95%), implying that negative findings are reliable in most cases. Interobserver agreement was slightly lower compared to the other modalities, yet still indicative of good overall agreement.

Previous studies have reported varying results regarding the diagnostic accuracy of TOF MRA for the follow-up of treated cerebral aneurysms.^8,16^ Specificity and PPV are generally high, often approaching 100%. Sensitivity, however, shows considerable variability, ranging from 50% in some studies to 83.3% in others.^8,16^ NPV is typically high as well, reported between 84% and 97.6%. These findings are broadly consistent with our results for pre-contrast TOF MRA in our cohort.

To our knowledge, the use of contrast enhanced TOF MRA sequence has not been previously studied. However, some research has explored contrast-enhanced MRA using different acquisition techniques.^7,16^ These studies also demonstrate variability, with reported sensitivity values ranging from 83% to 100%, and specificity generally high, with the lowest reported at 90.9% for contrast enhanced MRI.^7,8^ Although direct comparison with our results is limited due to methodological differences, the sensitivity observed in our post-contrast TOF MRA findings aligns with these previous studies. The specificity we observed, however, was slightly lower.

To demonstrate non-inferiority of the new imaging tests compared to DSA, at least 24 completely occluded aneurysms were required, allowing a maximum of one false negative. This threshold was based on power calculations for sCTA, as sensitivity data for TOF MRA were insufficient. In the sCTA group, 25 occlusions were included with 5 false negative observations. Pre-contrast TOF MRA had 4 false negatives among 24 occlusions. Only post-contrast TOF MRA met the non-inferiority requirement initially defined for sCTA with 24 occlusions and one false negative observation. The non-invasive nature of these imaging modalities offers a notable advantage. Although pre- and post-contrast TOF MRA were evaluated separately in this study, a combined approach may offer improved diagnostic performance by enhancing both sensitivity and specificity.

sCTA and TOF MRA offer a clear advantage over DSA: the ability to visualize the entire intracranial vasculature in a single acquisition. DSA is limited to the vessel selectively catheterized and a restricted number of adjacent vessels. This more comprehensive vascular overview provided by sCTA and TOF MRA enables the detection of aneurysms outside the initially targeted region. As a result, aneurysms in different vascular territories can be identified with these modalities, whereas they may remain undetected with DSA. This advantage proved clinically relevant in our cohort, where sCTA and TOF MRA facilitated the identification and subsequent treatment of two previously unrecognized aneurysms located in different vascular territories. Moreover, sCTA and TOF MRA were not associated with any complications in our study, whereas DSA-related complications such as puncture site hematomas, transient neurological deficit, contrast reaction or a thromboembolic event were observed in 5 cases. While most events were mild or transient, their occurrence underscores the procedural risks inherent to DSA, highlighting the advantages of sCTA and TOF MRA as safer alternatives.

This study has several limitations that should be acknowledged. First, we assumed DSA to be the reference standard with 100% accuracy. However, upon retrospective review, discrepancies were noted in two cases. In both, DSA indicated complete occlusion, whereas sCTA, pre- and post-contrast TOF MRA suggested incomplete occlusion. In one instance, the incomplete occlusion was retrospectively identifiable on DSA but had been scored as complete. In the other, assessment was hindered by coil overprojection, rendering the presence of residual flow uncertain.

Second, although the use of blinded image assessment minimized observer bias, it does not fully reflect routine clinical practice, in which prior imaging is often available to aid interpretation. In our study, observers did not have access to pre-treatment imaging. It is plausible that access to baseline scans would have facilitated post-treatment evaluation of aneurysm occlusion.

Finally, the use of an ultra-high-resolution CT scanner limits the generalizability of our findings, as such technology is not yet widely available in clinical settings.

Recent studies have emerged on the application of photon-counting detector CTA for the evaluation of treated and untreated intracranial aneurysms, offering high-resolution imaging at a reduced radiation dose compared to conventional CTA.^17–19^ Despite its currently limited availability in clinical setting, this imaging modality holds considerable promise for the evaluation of treated cerebral aneurysms.

## CONCLUSION

sCTA, pre- and post-contrast TOF MRA appear to be appropriate imaging modalities for the evaluation of aneurysmal occlusion after FDS placement, considering the high specificity and NPV. However, DSA should be considered in cases of diagnostic uncertainty, particularly when an aneurysm remnant is suspected, or when image quality is compromised by the presence of artefacts or other technical limitations.

## Data Availability

The data that support the findings of this study are available from the corresponding author upon request

## NON-STANDARD ABBREVIATIONS AND ACRONYMS

CI: confidence interval
DSA: digital subtraction angiography
FDS: flow diverter stent
MRA: MR angiography
NPV: negative predictive value
PPV: positive predictive value
RR: Raymond-Roy
SAH: subarachnoid hemorrhage
sCTA: subtraction CT angiography
SEMAR: single-energy metal artifact reduction
TOF: time-of-flight

